# Real-world utilization and initial experience with aflibercept-ayyh (PAVBLU^®^) for retinal disorders in United States retina practices: A descriptive retrospective analysis

**DOI:** 10.64898/2026.02.25.26345681

**Authors:** Argentina E. Servin, Ian McFadden, Houri Esmaeilkhanian, Diana Holcomb, James Lin, Carl C. Awh

**Affiliations:** Amgen Inc., 1 Amgen Center Drive, Thousand Oaks, CA; Retina Consultants of America, 601 State Street Suite 325, Southlake, TX 76092; Vitreoretinal Consultants of New York, 600 Northern Blvd # 216, Great Neck, NY, USA; Tennessee Retina, Centennial Professional Plaza, 345 23rd Avenue North, Ste. 350, Nashville, TN 37203

**Keywords:** Aflibercept-ayyh, age-related macular degeneration, anti-VEGF, ABP 938, biosimilar, diabetic retinopathy, diabetic macular edema, PAVBLU^®^, real world, retinal vein occlusion

## Abstract

**Introduction:** Anti-vascular endothelial growth factor (anti-VEGF) therapies are standards of care for vision-threatening retinal diseases. This retrospective observational study describes demographics, utilization, best recorded visual acuity (BRVA), and safety among eyes with neovascular age-related macular degeneration (nAMD), diabetic retinopathy (DR), diabetic macular edema (DME), or retinal vein occlusion (RVO) treated with the biosimilar aflibercept-ayyh (PAVBLU^®^) in routine clinical practice.

**Methods:** Electronic medical records from the Retina Consultants of America database of patients receiving aflibercept-ayyh (12/1/2024-10/31/2025) were analyzed, focusing on eyes with ≥84 days of follow-up. The index date was the first documented aflibercept-ayyh injection. Postindex data were used to assess treatment patterns, BRVA (Wilcoxon signed rank test), and adverse events of special interest (AESIs).

**Results:** A total of 1,000 consecutive eyes from 989 patients received 3,730 injections of aflibercept-ayyh; most (91%) switched from prior anti-VEGF therapy and 9% were anti-VEGF treatment-naïve. Disease distribution was 58% nAMD, 19% RVO, 16% DME, and 7% DR. Among switchers, median (IQR) number of prior injections was 21 (8–46). Median (IQR) follow-up was 6.0 months (4.6–7.1). Median (IQR) number of aflibercept-ayyh injections per eye was 4 (3–5). Among eyes with ≥84 days of follow-up (n=889), mean BRVA expressed as logarithm of minimum angle of resolution (logMAR) remained stable for switchers (0.4 to 0.4; *P*=0.96) and improved from baseline in anti-VEGF-naïve eyes (0.5 to 0.4; *P*<0.01). Confirmed AESIs included iritis (n=2; 0.05% of injections), with no events of vitreous cells, endophthalmitis, retinal detachment, retinal vasculitis, or vitreous hemorrhage.

**Conclusion:** In this descriptive real-world analysis, aflibercept-ayyh was associated with stable visual acuity in previously treated eyes and vision improvement in treatment-naïve eyes, with no new or unexpected safety findings, consistent with expectations for aflibercept. These findings add real-world experience to preexisting evidence demonstrating no clinically meaningful differences between aflibercept-ayyh (PAVBLU^®^) and reference aflibercept (EYLEA^®^).

**KEY SUMMARY POINTS:** *Why carry out this study?:*

- The anti–vascular endothelial growth factor (VEGF) drug aflibercept, approved in 2011 and marketed in the United States as EYLEA^®^,* has demonstrated efficacy in treating retinal diseases such as neovascular age-related macular degeneration (nAMD), diabetic retinopathy (DR), diabetic macular edema (DME), or retinal vein occlusion (RVO) and is a standard of care for these disorders.
- Aflibercept-ayyh is a biosimilar to aflibercept that has demonstrated comparable efficacy and safety in the treatment of nAMD in a randomized controlled clinical trial.
- This study describes the real-world use patterns, vision outcomes, and safety of aflibercept-ayyh in clinical settings in the United States for the treatment of nAMD, DR, DME, and RVO.

*What was learned from the study?:*

- In this real-world study of 1,000 consecutive eyes treated with the biosimilar aflibercept-ayyh in patients with retinal diseases, we observed no new safety concerns and that aflibercept-ayyh maintained visual acuity in eyes switching anti-VEGF agents and improved vision in anti-VEGF–naïve eyes, consistent with expected responses to aflibercept.
- These findings support aflibercept-ayyh as a suitable treatment option when anti-VEGF therapy is indicated. *EYLEA^®^ is a registered trademark of Regeneron Pharmaceuticals, Inc. PAVBLU^®^ is a registered trademark of Amgen Inc.

## INTRODUCTION

Neovascular age-related macular degeneration (nAMD), diabetic retinopathy (DR), diabetic macular edema (DME), and retinal vein occlusion (RVO) are leading causes of vision impairment and blindness worldwide.^1^ These diseases are characterized by vascular dysfunction, including pathological neovascularization and/or increased vascular permeability, which can result in progressive vision loss if left untreated.^2–5^ Over the past two decades, anti–vascular endothelial growth factor (anti-VEGF) therapies such as bevacizumab, ranibizumab, and aflibercept have advanced the management of retinal diseases^1^ and are now established as the standard of care across these indications.^6^

Aflibercept, marketed in the United States as EYLEA^®^, has demonstrated efficacy and an established safety profile in the treatment of nAMD,^7^ DR and DME,^8^ and RVO.^9, 10^ Aflibercept-ayyh (PAVBLU^®^) is a biosimilar to aflibercept approved by the United States Food and Drug Administration (FDA) for these four indications.^11^ As with all biosimilars, regulatory approval required a comprehensive demonstration of no clinically meaningful differences to the reference product, supported by analytical, functional, nonclinical, and clinical evidence. In a randomized controlled trial in patients with nAMD, aflibercept-ayyh demonstrated efficacy and safety comparable to that of reference aflibercept, with additional analytical and functional studies confirming structural and biological similarity.^12–14^

Real-world evidence is important for understanding how therapies perform in routine clinical practice, particularly across diverse patient populations and treatment histories. To date, limited real-world data are available describing utilization patterns, visual outcomes, and safety of aflibercept-ayyh following its introduction into clinical practice. This retrospective observational study aims to describe the real-world use, patient outcomes, and safety profile of aflibercept-ayyh among patients with nAMD, DR, DME, and RVO treated in United States retina practices.

## METHODS

### Study Design and Data Source

This descriptive, retrospective, observational analysis used electronic medical records (EMRs) from Retina Consultants of America (RCA), a retina-only eye care specialist network, to evaluate real-world treatment patterns, visual outcomes, and safety among eyes treated with aflibercept-ayyh in the United States. Data were extracted from EMRs of patients seen by 175 retina specialists at 146 locations across 24 states using Structured Query Language (SQL). Records from December 1, 2024, to October 31, 2025, were included in this analysis.

### Ethics/Ethical Approval

The study was reviewed by an independent external institutional review board (IRB), Advarra, and was determined to be exempt from IRB oversight (determination date: June 17, 2025) due to its use of de-identified data collected during routine clinical care. Informed consent was not required. The study was conducted in accordance with applicable regulatory requirements and the principles of the Declaration of Helsinki.

### Study Population

Eligible eyes were from adult patients (≥18 years old) with a documented diagnosis of nAMD, DR, DME, or RVO who received at least one intravitreal injection of aflibercept-ayyh during the study period. A best recorded visual acuity (BRVA) of 20/400 or better was required for study inclusion of each eye.

The index date was defined for each eye as the date of the first documented aflibercept-ayyh injection. Analyses of BRVA outcomes focused on eyes with at least 84 days of follow-up after the index date, corresponding to the planned primary assessment time point of approximately 3 months. Descriptive summaries of treatment utilization and safety included all eligible eyes with available postindex data. The first 1,000 consecutively treated eyes meeting eligibility criteria were included. Bilaterally treated eyes from the same patient were included and analyzed as independent study eyes.

### Outcomes

Patient demographics (age, sex, race and ethnicity, and smoking status) were summarized as reported by the patient using values recorded at the index date. All other outcomes were summarized by study eye. For all analyses, eyes were categorized into two cohorts by baseline status: (1) Anti-VEGF treatment-naïve eyes and (2) Eyes that switched to aflibercept-ayyh after prior treatment with another anti-VEGF agent. Some outcomes were analyzed in a subset of eyes that had at least one follow-up visit and EMR data available with follow-up of at least 84 days (∼3 months) from the first injection of aflibercept-ayyh.

Baseline clinical characteristics were identified using clinical documentation as recorded in EMRs and relevant International Classification of Diseases, Tenth Revision (ICD-10), codes. Treatment patterns were characterized using Healthcare Common Procedure Coding System [HCPCS]) Q-codes and procedure codes to identify administered anti-VEGF agents and doses. The number of previous anti-VEGF injections received per eye as documented in EMRs was also extracted. The injection interval was defined as the number of days between consecutive anti-VEGF injections.

BRVA documented as standard Snellen fractions was extracted and converted to logarithm of the minimum angle of resolution (logMAR) units to facilitate statistical analysis.

Adverse events of special interest (AESIs), defined as adverse events occurring within 21 days of any aflibercept-ayyh treatment and not existing at the time of injection,^15, 16^ were identified in EMRs according to clinical and imaging findings and supported by ICD-10 codes, with manual chart reviews performed by investigating physicians to validate extracted data and adjudicate ambiguous or missing cases. Defined AESIs were endophthalmitis, retinal vasculitis, iritis, vitreous cells, retinal detachment, and vitreous hemorrhage.

Information was extracted from the EMR system through direct queries of structured data fields. When necessary, additional data were abstracted manually by investigators or qualified team members through detailed review of treatment records. When needed, diagnoses identified using standardized ICD-10 codes were verified through supporting clinical documentation. For data elements not captured in structured EMR fields, manual chart reviews were conducted. All abstractions were conducted by trained and experienced personnel following consistent procedures to enhance data quality and reproducibility. All data were collected during routine clinical care, and there was no direct contact with patients.

### Statistical Analysis

Descriptive statistics were used to summarize patient demographics, baseline clinical characteristics, treatment utilization, visual acuity outcomes, and safety events. Continuous variables were summarized using means and standard deviations or medians and interquartile ranges (IQRs), as appropriate, and categorical variables were summarized using frequencies and percentages.

Within-eye changes in BRVA (expressed as logMAR units) from baseline to follow-up were evaluated using the Wilcoxon signed rank test for paired data. These analyses were performed overall and separately for treatment-naïve and switcher cohorts, as well as by primary diagnosis. *P*-values were reported to describe the direction and magnitude of observed within-eye changes.

Incidences of AESIs were summarized descriptively using event counts and relevant exposure denominators, including the total number of aflibercept-ayyh injections. AESIs were identified within 21 days following an aflibercept-ayyh injection and adjudicated through manual chart review to confirm event occurrence and timing. Given the low frequency of events and the descriptive nature of the analysis, safety outcomes were not intended for formal hypothesis testing.

All analyses were conducted using Python version 3.13.0.

## RESULTS

### Study Population

Between December 1, 2024, and October 31, 2025, the first 1,000 consecutively treated eyes meeting eligibility criteria belonging to 989 patients received at least one aflibercept-ayyh injection. Among patients, median age was 79 years, 55% were female, and 63% were never smokers. The linked diagnoses were nAMD, RVO, DME, and DR in 578 (58%), 193 (19%),162 (16%), and 67 (7%) eyes, respectively. Of these eyes, 906 (91%) had previously been treated with at least one other anti-VEGF agent (switchers) and 94 (9%) were anti–VEGF treatment naïve. Of the 906 eyes that had switched, median (IQR) number of prior injections was 21 (8–46). The most frequent prior anti-VEGF therapies among evaluable patients who switched were aflibercept (327 [39%] eyes) and/or ranibizumab-eqrn (Cimerli, 215 [26%] eyes) (Table 1). At the time of the analysis (data cutoff: October 31, 2025), median (IQR) follow-up per study eye was 6.0 (4.6–7.1) months and 922 (92%) eyes had at least one follow-up visit and EMR data available with follow-up of at least 84 days (∼3 months) from the first injection of aflibercept-ayyh (Figure 1).

**Figure 1.**
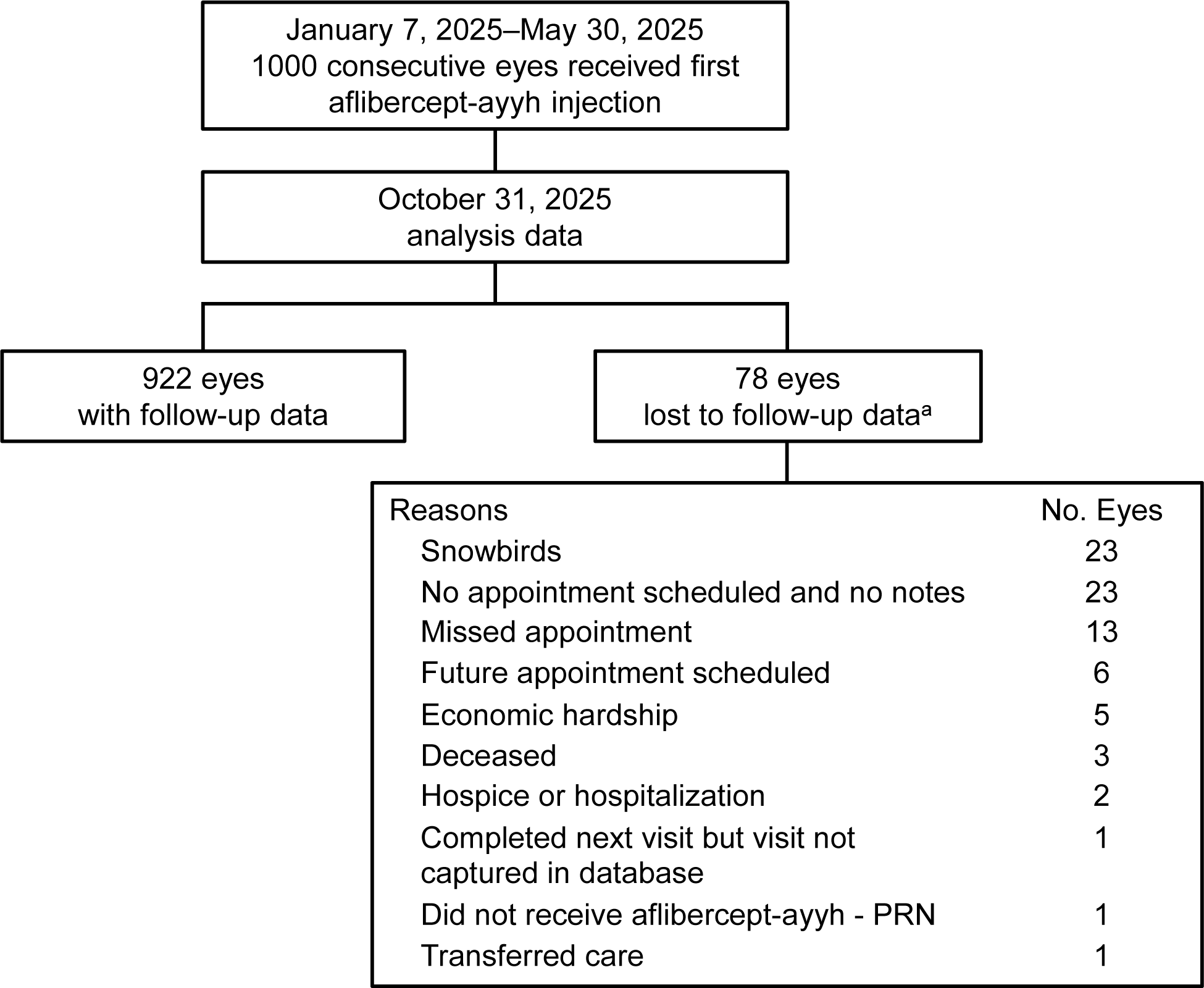
Study eye disposition ^a^ Reasons for 78 eyes not having ≥84 days of follow-up at time of data cutoff (October 31, 2025). Study eye flow throughout the study is shown, including reasons for lost to follow-up. PRN, pro re nata, ie, as needed.

**Table 1.** Demographic and clinical parameters.

|  | **Treatment Naïve** | **Switchers** | **All** |
| --- | --- | --- | --- |
| Patients, n | 94 | 895 | 989 |
| Both eyes | 0 | 11 | 11 |
| Eyes, no. | 94 | 906 | 1,000 |
| **Patients** |  |  |  |
| Age, years |  |  |  |
| Mean (SD) | 78 (10) | 78 (12) | 78 (11) |
| Median (IQR) | 78 (72–84) | 79 (72–87) | 79 (72–86) |
| Sex, n (%) |  |  |  |
| Female | 54 (57) | 493 (55) | 547 (55) |
| Male | 37 (39) | 385 (43) | 422 (43) |
| Unknown | 3 (3) | 17 (2) | 20 (2) |
| Smoking status |  |  |  |
| Never smoker | 50 (53) | 573 (64) | 623 (63) |
| Former smoker | 39 (41) | 272 (30) | 311 (31) |
| Current smoker | 5 (5) | 50 (6) | 55 (6) |
| **Eyes** |  |  |  |
| Diagnosis, no. (%) |  |  |  |
| nAMD<sup>a</sup> | 58 (62) | 520 (57) | 578 (58) |
| DR<sup>b</sup> | 9 (10) | 58 (6) | 67 (7) |
| DME | 12 (13) | 150 (17) | 162 (16) |
| RVO<sup>c</sup> | 15 (16) | 178 (20) | 193 (19) |
| Prior injections, no. |  |  |  |
| Mean (SD) | 0 | 29 (26) | 29 (26) |
| Median (IQR) [range] | 0 | 21 (8–46) [1–112] | 21 (8–46) [1–112] |
| Prior anti-VEGF therapies, no. |  |  |  |
| Mean (SD) | 0 | 1.2 (0.6) | 1.2 (0.7) |
| Median (IQR) [range] | 0 | 1 (1–2) [0–3] | 1 (1–1) [0–3] |
| Prior anti-VEGF therapy, no. (%)<sup>d</sup> |  |  |  |
| Aflibercept | 0 | 329 (39) | 329 (33) |
| Ranibizumab-eqrn (Cimerli) | 0 | 215 (25) | 215 (22) |
| Ranibizumab | 0 | 26 (3) | 26 (3) |
| Bevacizumab | 0 | 30 (4) | 30 (3) |
| Faricimab | 0 | 31 (4) | 31 (3) |
| Not identified | 0 | 214 (25) | 214 (21) |

<sup>a</sup>nAMD=nAMD with active CNV, nAMD with inactive CNV, and nAMD unspecified.
<sup>b</sup>DR=PDR, moderate PDR, and mild PDR.
<sup>c</sup>RVO=branch RVO and central RVO.
<sup>d</sup>Denominator of 845 for switcher eyes column.
Prior anti-VEGF therapy data were not available for all switcher eyes as some received care outside of the RCA network (ie, numbers will not add up to the total number of eyes).
CNV, choroidal neovascularization; DME, diabetic macular edema; DR, diabetic retinopathy; IQR, interquartile range; nAMD, neovascular age-related macular degeneration; PDR, proliferative diabetic
retinopathy; RCA, Retina Consultants of America; RVO, retinal vein occlusion; SD, standard deviation; VEGF, vascular endothelial growth factor.

### Treatment

A total of 3,730 injections were administered to study eyes during the study period. Median (IQR) number of aflibercept-ayyh injections per eye was 4 (3–5), with a median (IQR) injection interval of 54 (35–70) days. For switchers and naïve eyes, median injection intervals were 56 and 37 days, respectively (Table 2).

**Table 2.** Aflibercept injections.

|  | **Treatment Naïve** | **Switchers** | **All** |
| --- | --- | --- | --- |
| Eyes, no. | 94 | 906 | 1,000 |
| Eyes with follow-up data | 77 | 845 | 922 |
| Injection interval, days |  |  |  |
| Mean (SD) | 44 (20) | 59 (26) | 57 (26) |
| Median (IQR) [range] | 37 (33–49) [26–203] | 56 (42–70) [28–278] | 54 (35–70) [28–278] |
| Total injections, no. | 395 | 3,335 | 3,730 |
| Mean (SD) | 4.20 (2.0) | 3.68 (1.7) | 3.73 (1.7) |
| Median (IQR) [range] | 4 (2–6) [1–9] | 3 (3–5) [1–11] | 4 (3–5) [1–11] |
| Aflibercept-ayyh injections,<br>no. (%) |  |  |  |
| 1 | 94 (100) | 906 (100) | 1,000 (100) |
| 2 | 85 (90.4) | 828 (91.5) | 913 (91.3) |
| 3 | 70 (74.5) | 689 (76.0) | 759 (75.9) |
| 4 | 59 (62.8) | 443 (48.9) | 502 (50.2) |
| 5 | 45 (47.9) | 252 (27.8) | 297 (29.7) |
| 6 | 25 (26.6) | 117 (12.9) | 142 (14.2) |
| 7 | 10 (10.6) | 55 (6.1) | 65 (6.5) |
| 8 | 5 (5.3) | 29 (3.2) | 34 (3.4) |
| ≥9 | 2 (2.1) | 16 (1.8) | 18 (1.8) |
| Injections per eye |  |  |  |
| nAMD, no. of eyes |  |  |  |
| Mean (SD) | 4.6 (1.9) | 3.7 (1.7) | 3.8 (1.7) |
| Median (IQR) | 5 (3–6) | 3 (3–5) | 4 (3–5) |
| DR, no. of eyes |  |  |  |
| Mean (SD) | 3.1 (2.0) | 3.1 (1.7) | 3.1 (1.7) |
| Median (IQR) | 2.5 (1.5–5) | 3 (2–4) | 3 (2–4) |
| DME, no. of eyes |  |  |  |
| Mean (SD) | 4.2 (2.4) | 3.5 (1.7) | 3.6 (1.7) |
| Median (IQR) | 4 (2–5.5) | 3 (2–5) | 3 (2–5) |
| RVO, no. of eyes |  |  |  |
| Mean (SD) | 3.5 (1.7) | 4.0 (1.7) | 4.0 (1.7) |
| Median (IQR) | 4 (2–5) | 4 (3–5) | 4 (3–5) |

Numbers may add up to less than the number of eyes injected due to missing data.
DME, diabetic macular edema; DR, diabetic retinopathy; IQR, interquartile range; nAMD, neovascular age-related macular degeneration; RVO, retinal vein occlusion; SD, standard deviation.

### Visual Acuity

In eyes of patients who switched from a prior anti-VEGF therapy, had 3-month follow-up data, and were assessable for visual acuity (n=815), there was no change from baseline in mean BRVA observed since beginning aflibercept-ayyh treatment (logMAR 0.4 to 0.4, *P*=0.96, Table 3). In eyes of patients who were anti-VEGF–naïve (n=74), vision significantly improved from mean logMAR 0.5 to 0.4 (*P*<0.01).

**Table 3.** BRVA after aflibercept-ayyh injection.

| **LogMAR Value (BRVA)** | **Baseline** | **≥3 Months** | ***P* Value<sup>a</sup>** |
| --- | --- | --- | --- |
| **Treatment naïve** |  |  |  |
| No. of eyes with logMAR value | 74 | 74 |  |
| Mean (SD) | 0.5 (0.5) | 0.4 (0.5) | <0.01 |
| Median (IQR) | 0.4 (0.2–0.6) | 0.3 (0.1–0.5) |  |
| **Switchers** |  |  |  |
| No. of eyes with logMAR value | 815 | 815 |  |
| Mean (SD) | 0.4 (0.5) | 0.4 (0.5) | 0.96 |
| Median (IQR) | 0.3 (0.2–0.6) | 0.3 (0.1–0.5) |  |

<sup>a</sup>Wilcoxon signed rank test.
Baseline vision as recorded before the first aflibercept-ayyh injection.
BRVA, best recorded visual acuity; IQR, interquartile range; logMAR, logarithm of minimum angle of resolution; SD, standard deviation.

Similar results with respect to changes in visual acuity were observed across diagnosis subgroups. Median baseline BRVA according to diagnosis ranged from 0.40 to 0.18 logMAR units for switchers and median change from baseline was 0 for each diagnosis subgroup. Anti-VEGF–naïve eyes were observed to have a baseline BRVA of 0.54–0.24 logMAR and showed directional improvement across all diagnoses among eyes with at least 3 months of follow-up (–0.08 to –0.19), albeit in smaller sample sizes (Figure 2).

**Figure 2.**
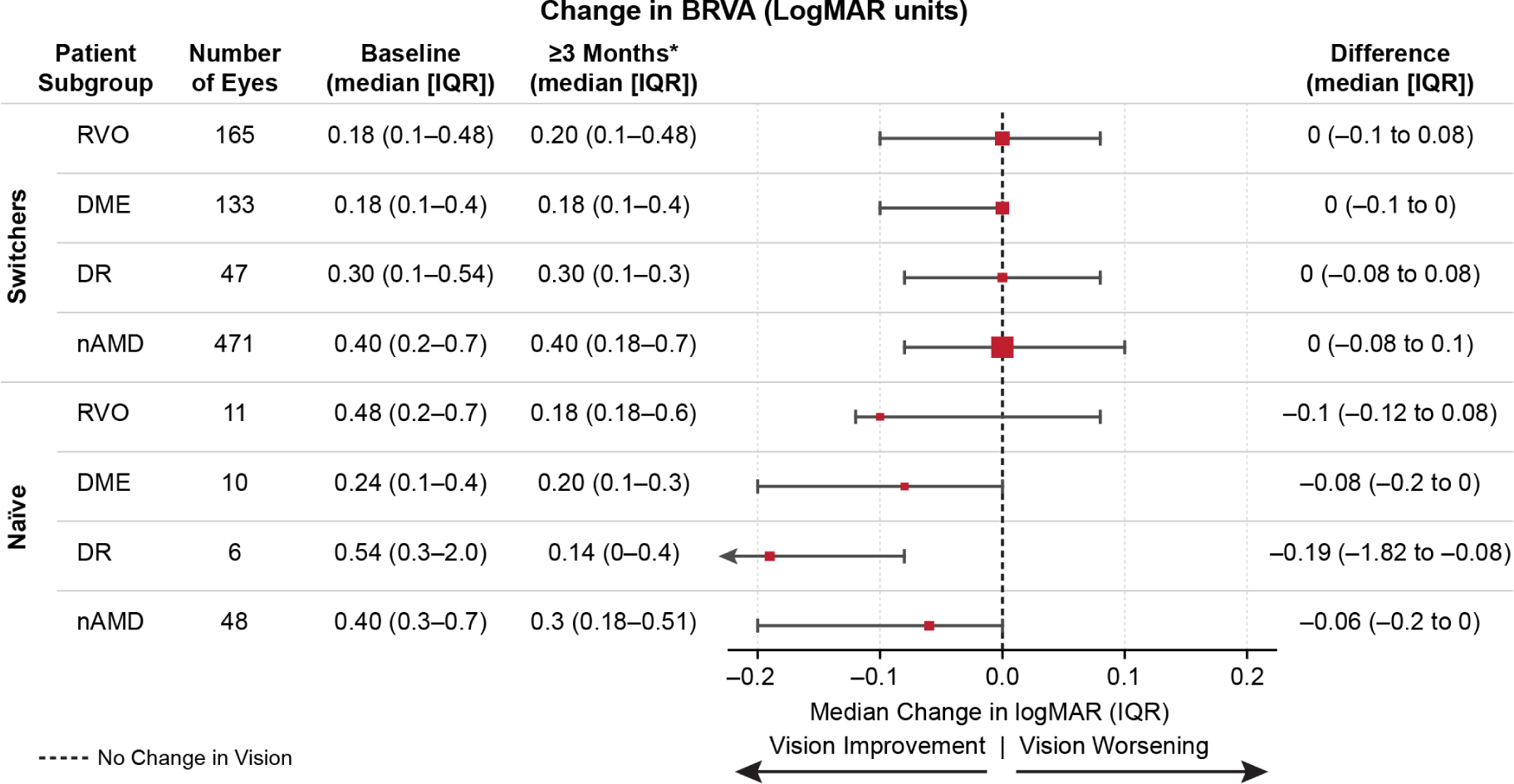
Change in BRVA *With a minimum of 84 days of follow-up. Change in BRVA from baseline to ≥3 months of follow-up is shown by cohort (switcher or treatment-naïve) and disease. Disease categories were defined as follows: nAMD consists of nAMD with active CNV, nAMD with inactive CNV, and nAMD unspecified; DR consists of PDR, moderate PDR, and mild PDR; and RVO consists of branch RVO and central RVO. CNV, choroidal neovascularization; DR, diabetic retinopathy; IQR, interquartile range; logMAR, logarithm of the minimum angle of resolution; nAMD, neovascular age-related macular degeneration; PDR, proliferative diabetic retinopathy; RVO, retinal vein occlusion.

### Safety Analysis

There were two confirmed events of iritis, and no events of vitreous cells, endophthalmitis, retinal detachment, retinal vasculitis, or vitreous hemorrhage occurred following aflibercept-ayyh treatment. In one of the two cases of iritis, the contralateral eye previously also experienced iritis following injection of a different anti-VEGF agent. Both cases of iritis resolved. Among 3,730 aflibercept-ayyh injections in this analysis, the incidence rate for iritis was 0.54 events per 1,000 injections (0.05%).

## DISCUSSION

Anti-VEGF therapies remain the longstanding standard of care for nAMD, DME, DR, and RVO, and biosimilars can provide new options for patients, physicians, and the broader healthcare system. Rigorous regulatory pathways ensure that biosimilars show structural and functional similarity to reference biologics, and that no clinically meaningful differences in safety, efficacy, or immunogenicity are demonstrated. Nonetheless, real-world evidence is essential to demonstrate that patient outcomes in routine clinical practice remain consistent with the known benefit-risk profiles of the reference medicines and to help characterize the potential for rarely occurring events or adverse reactions.^17–21^

This real-world analysis provided insight into the use of aflibercept-ayyh in routine clinical practice. In the first 1,000 eyes treated consecutively across the RCA practice network, the utilization patterns aligned with current recommendations for the treatment of retina disorders.^22^ As expected, dosing intervals were shorter in treatment-naïve eyes vs eyes that switched from another anti-VEGF agent, consistent with naïve eyes receiving monthly anti-VEGF loading doses and switching eyes receiving extended maintenance dosing.^23^ After 3 or more months of follow-up, visual acuity improved in treatment-naïve eyes and remained stable in eyes that switched, aligning with previously reported real-world outcomes for aflibercept.^24, 25^ These observations were directionally consistent across diagnoses for both cohorts, although small sample sizes in diagnosis subgroups of anti-VEGF–naïve eyes limit interpretability.

Interpretation of visual acuity outcomes should be considered in the context of the study population and follow-up duration. The majority of eyes had extensive prior exposure to anti-VEGF therapy, reflecting chronic disease and limited potential for short-term visual improvement. In this setting, maintenance of visual acuity over the early follow-up period is consistent with treatment goals and aligns with previously reported real-world outcomes following switches between anti-VEGF agents.^23–25^ In contrast, the observed improvement among treatment-naïve eyes is consistent with expected responses during the initial treatment phase of anti-VEGF therapy. The findings with aflibercept-ayyh in this analysis are aligned with real-world observations of improvement in visual acuity of anti-VEGF treatment-naïve eyes treated with aflibercept (EYLEA^®^) for nAMD and maintenance of visual acuity in eyes switching from a prior anti-VEGF to aflibercept (EYLEA^®^) for the treatment of wet AMD or DME.^23–25^

Importantly, no new or unexpected safety signals emerged during the observation period and the safety profile observed was consistent with historical experience for aflibercept and other intravitreal anti-VEGF agents in real-world settings.^26, 27^ Following 3,730 intravitreal injections of aflibercept-ayyh, two cases of iritis were reported. Of note, one case of iritis subsequently followed a previously reported case in the contralateral eye after injection of a different anti-VEGF agent. Both cases of iritis resolved with topical steroid treatment and neither sustained permanent visual loss. While the sample size in this analysis is too small to approximate a definitive rate, the observations are consistent with iritis/uveitis/intraocular inflammation rates historically observed with aflibercept (EYLEA^®^).^11, 28, 29^ These findings and the low incidence of AESIs overall are consistent with the expected rates of iritis and known inflammatory events associated with intravitreal anti-VEGF injections.^30^

This study has several limitations inherent to its retrospective, observational design. The absence of randomization and a concurrent comparator group preclude direct comparisons of outcomes between aflibercept-ayyh and reference aflibercept or other anti-VEGF therapies. While no unexpected outcomes were observed in the study period, follow-up duration was relatively short, limiting assessment of longer-term visual outcomes, durability of response, and rare AESIs. Continued follow-up from this cohort and additional real-world studies will be conducted to further characterize the long-term outcomes, durability of response, and safety of aflibercept-ayyh as its clinical use expands. Additionally, reliance on routinely collected EMR data may introduce limitations or bias due to incomplete or missing information. For example, reasons for switching to aflibercept-ayyh were not systematically captured, limiting interpretation of treatment decisions and subgroup analyses. Although a small proportion of patients contributed bilateral eyes (n=11), each eye was analyzed independently, which may introduce minor correlation bias. There was also a preponderance of eyes that switched (n=906) and fewer treatment-naïve eyes (n=94), limiting interpretation of both safety and visual acuity outcomes among anti-VEGF–naïve eyes, especially within subgroups by primary diagnosis. Finally, anatomical outcomes such as optical coherence tomography measures were not evaluated in this analysis.

Despite these limitations, the study has notable strengths, including its large overall sample size, broad geographic representation, inclusion of multiple retinal indications, and use of consecutively treated eyes, which together enhance the generalizability of the findings to routine retinal practice in the United States. Physician-adjudicated validation of AESIs further strengthens the reliability of the safety findings.

## CONCLUSION

This descriptive, retrospective analysis of 1,000 consecutively treated eyes from retina practices across the United States provides real-world insight into the clinical use of aflibercept-ayyh. Visual acuity was maintained in eyes that switched from prior anti-VEGF therapy and improved in anti-VEGF–naïve eyes, consistent with real-world expectations for aflibercept (EYLEA^®^), and no new or unexpected safety findings were observed during the observation period.

These findings add to the existing body of evidence established in analytical and clinical studies demonstrating no clinically meaningful differences between aflibercept-ayyh (PAVBLU^®^) and reference aflibercept (EYLEA^®^).

## Medical Writing, Editorial, and Other Assistance

Medical writing assistance was provided by Julie Gegner, PhD, and Susanna Mac, MD, PhD, both funded by Amgen. Biostatistical programming support was provided by Kruti Doshi, Patrick Ventura, Nalini Kethineni, and Baitong Wang of Amgen Inc.

## Authorship Contributions

Study design: Argentina E. Servin, Houri Esmaeilkhanian, Diana Holcomb, James Lin, and Carl C. Awh; data collection: Houri Esmaeilkhanian, Diana Holcomb, James Lin, and Carl C. Awh; data analysis and critical review of the manuscript: all authors.

## Data Availability

The data supporting the findings of this study are derived from electronic medical records from Retina Consultants of America and are not publicly available due to patient privacy protections and contractual restrictions governing access to these data. De-identified data may be made available from the corresponding author upon reasonable request, subject to approval by Retina Consultants of America and execution of appropriate data use agreements.

